# Cost-effectiveness of generic semaglutide for Status First Nations adults with obesity and prediabetes under British Columbia’s Plan W

**DOI:** 10.64898/2026.11.24.26368896

**Authors:** Thomas E. Curry, Ilya Kovalko, Jessica E. Stockdale, Eldon Spackman

**Author notes:** Corresponding author: Thomas E. Curry, Indigenous Health, Fraser Health Authority, Surrey, British Columbia, Canada.

## Abstract

**Background:** British Columbia’s Plan W could cover generic semaglutide for Status First Nations adults with obesity and prediabetes. Its value for money is unknown.

**Methods:** We modelled lifetime public costs and quality-adjusted life-years for adults with obesity and hemoglobin A1c of 6.0% to 6.4% who added semaglutide 1 mg to usual care, using the best available Canadian diabetes costs (Alberta and Ontario). After stopping, everyone returned to usual diabetes risk. In addition, the cases avoided on treatment either stayed avoided (100%), mostly stayed avoided (83%, from one-year trial data), or all occurred within a year (0%).

**Results:** At 83%, treatment gained 0.124 quality-adjusted life-years and cost $4,250 (Alberta) or $16,297 (Ontario-derived) per quality-adjusted life-year. At 0%, costs were $39,500 and $46,376. At 100% with Alberta costs, spending fell by $38.

**Conclusions:** Generic semaglutide is likely a cost-effective way to postpone diabetes. Whether it pays for itself is uncertain.

## Introduction

For an adult with obesity and a hemoglobin A1c (HbA1c) of 6.0% to 6.4%, progression to diabetes changes both the care required and the years spent living with the disease. Lower-priced generic semaglutide raises a coverage question for British Columbia’s Plan W, which pays for eligible prescriptions across adulthood for Status First Nations people in BC whose eligibility is confirmed by the First Nations Health Authority. Would funding treatment before diabetes develops yield enough health gain to justify the additional public spending?[1]

In the SELECT trial, semaglutide reduced progression to diabetes-range HbA1c among participants with overweight or obesity and established cardiovascular disease. The treatment effect was apparent in participants with higher baseline HbA1c. Extending this finding to prevention in Plan W requires several judgments. The dose evaluated in SELECT was 2.4 mg, many patients stop treatment, and our population of interest has no established cardiovascular disease. The economic return also depends on the cost of care while living with diabetes. Two Canadian studies provide different estimates of that cost.[2-6]

We estimated the lifetime health gains and costs of adding semaglutide 1 mg to usual care for Plan W adults with obesity and higher-risk prediabetes. We applied the two Canadian cost sources to the same clinical course. This allowed us to distinguish the effect of postponing diabetes from the unit cost assigned to the resource use avoided.

## Methods

### Population, intervention, and perspective

The eligibility criterion was body mass index (BMI) of 30 kg/m^2^ or more and HbA1c 6.0% to 6.4%, without diagnosed diabetes or established cardiovascular disease. The modelled cohort entered at age 50 with a representative weight of 95 kg and height of 1.68 m (BMI 33.7 kg/m2). The cohort was chosen to represent Plan W adults who would meet the criterion; no Plan W data on their age or weight were available. Usual care included lifestyle support and indicated cardiovascular risk management. The intervention added subcutaneous semaglutide titrated to 1 mg weekly.

The Canadian indication for generic semaglutide that references Ozempic is type 2 diabetes. The use modelled here, in adults without diabetes, would be off-label. Semaglutide 2.4 mg (Wegovy) has a separate weight management indication. Product approval does not establish Plan W reimbursement for this proposed use.[7-9]

Following the recommendations of Canada’s Drug Agency, we included costs borne by the public health system, including prescriptions, monitoring, and hospital care. We excluded out-of-pocket costs, unpaid time, and productivity. Costs and quality-adjusted life-years (QALYs) were discounted at 1.5% per year. Monetary values are in projected 2026 Canadian dollars. Reporting follows the CHEERS 2022 statement.[10-12]

### Clinical model and treatment exposure

An annual-cycle cohort model followed patients until death or age 100. Patients could remain free of a new major event, experience myocardial infarction or stroke, develop kidney failure, or die. Within the surviving cohort, the model tracked diabetes and its contribution to cardiovascular events, kidney failure, costs, and quality of life. QALYs incorporated time in each health state, diabetes, weight change, and early adverse effects. Background mortality was multiplied by 1.5 (range 1.2 to 1.85) to reflect higher all-cause mortality among Status First Nations people in BC. The supplement describes the transition calculations.[13-16]

No local estimate existed for this BMI and HbA1c subgroup, so we assumed an untreated annual diabetes hazard of 0.062. This is the constant rate that would give 17% with diabetes after three years, the proportion of placebo participants in the higher-HbA1c subgroup of SELECT who had diabetes-range HbA1c at week 156. SELECT enrolled people with cardiovascular disease, and that proportion describes status at a single visit, so we treated the value as an assumption and tested alternatives. In the same subgroup, the hazard ratio for a first HbA1c of 6.5% or higher was 0.23. Because 1 mg produced 81% of the placebo-adjusted HbA1c reduction of 2.4 mg in the BELIEVE trial, we multiplied the log hazard ratio by 0.81. At full exposure, the resulting hazard ratio of about 0.30 lowers annual diabetes risk from about 6.0% to 1.8%. This estimate extrapolates across doses and from an HbA1c threshold to clinical diabetes. We assumed no direct cardiovascular benefit of semaglutide in primary prevention.[2,3,17]

We assumed 65% of people starting treatment remained on it at one year (range 47% to 80%) and 80% of continuing patients remained each subsequent year. Persistence under full public coverage has not been measured in Plan W. It was lower in United States practice, where 35% of adults without diabetes remained on treatment at one year and 16% at two years.[4] We tested those values in a sensitivity analysis. People who stopped accrued treatment and monitoring costs until the midpoint of their stopping year. At the point estimates this gave 0.825 years of exposure per person in year one before mortality adjustment. On-treatment effects reflected the same exposure, scaled to an assumed trial exposure of 85% and capped at the trial effect.

### Diabetes after treatment ends

Patients who continue treatment keep its on-treatment effects. Patients who stop lose the on-treatment reduction in diabetes risk but may stop with fewer diagnoses than comparable usual-care patients. We modelled three assumptions about what happens to this gap. With 100% retention, usual incidence resumes with no withdrawal-related rise in diagnoses, although the gap may narrow because more treated patients remain at risk. With 83% retention, 17% of the gap closes in the year after stopping.[18] With 0% retention, which we call complete catch-up, the whole gap closes over that year. After that first year, none of the three scenarios assumes any further catch-up from stopping. Whatever gap remains lasts for life and narrows only as both groups go on developing diabetes at the usual rate. All three scenarios count the diabetes-free time accrued before stopping. Weight-related quality of life followed the same post-treatment course in all three.

The intermediate value of 83% comes from trial data, not from the model. In Figure 3 of the STEP 1 extension, which followed participants for one year after they stopped semaglutide 2.4 mg, 0 of 141 semaglutide recipients and 3 of 53 placebo recipients with prediabetes at baseline had diabetes-range HbA1c (6.5% or more) at week 68. One year after withdrawal, the counts were 2 of 120 and 3 of 47.[18] The difference between groups narrowed from 5.66 to 4.72 percentage points, a ratio of 0.83. We call this the STEP 1 diabetes-range contrast. The estimate rests on very few events and on the participants who remained at follow-up. The ratio therefore describes the observed difference in prevalence. It cannot identify the incidence of confirmed diabetes after stopping 1 mg or separate withdrawal from ordinary progression.

### Public costs

The BC product payment used in the model was $84.39 per four-dose pen. Thirteen pens plus four $11 dispensing fees gave $1,141.07 per full treatment-year. We allowed $243 annually for incremental monitoring, prorated to time on treatment. We valued acute cardiovascular events separately. Published costs were converted to 2026 dollars using the Canadian health care price index, with a 2% projection from 2025.[12,19-21]

We used two estimates of the annual cost of diabetes. The Alberta estimate was $2,726, derived from the $2,352 cost in 2021 reported by Olivieri and colleagues. Its microvascular complications category includes pharmacy. The Ontario-derived estimate was $1,646, based on the $1,459 cost in 2022 used by Wiggins and colleagues. That study drew on modelled costs and Ontario attributable-cost data. Neither source identifies paid claims specifically for Plan W adults meeting the criterion. We did not add a separate prescription allowance because some drug expenditure is already included and the omitted share is unknown.[5,6,22]

### Analysis

For each of 10,000 draws we sampled uncertain parameters, evaluated both treatment arms with those inputs, and calculated the paired cost and QALY differences. Both cost specifications used identical clinical draws. Supplement Table S5 lists each sampled parameter with its distribution, range, and source. The 95% uncertainty intervals (UIs) are the 2.5th and 97.5th percentiles of the paired differences within each specified post-treatment course. Cost per QALY is the ratio of mean incremental cost to mean incremental QALYs. We calculated the probability that the additional health was worth the additional spending across a range of values per QALY. The $50,000 benchmark is illustrative.[10]

We examined lower and higher untreated diabetes hazards (0.03 and 0.10 per year), a four-year limit on new diabetes and weight-related benefits, the lower persistence observed in United States practice, and the Ontario formulary comparator price of $78.141 per pen. For each withdrawal course we calculated the pen price at which mean incremental spending reached zero. The model was written in Python. The code, inputs, and results are provided with the supplement.

## Results

### Health gains and lifetime public spending

With full retention, semaglutide gained 0.131 QALYs per person starting treatment (95% UI 0.047 to 0.261). Mean lifetime spending fell by $38 using Alberta costs (95% UI −$2,221 to $2,436) and rose by $1,633 using Ontario-derived costs (95% UI −$361 to $4,265). Spending fell in 53.2% of draws with Alberta costs and 5.5% with Ontario-derived costs. On average, semaglutide was cost-saving with Alberta costs and cost $12,424 per QALY with Ontario-derived costs. At $50,000 per QALY, the probability of positive net benefit was 99.8% with Alberta costs and 98.6% with Ontario-derived costs (Table 2, Figures 1 and 2).

**Table 1.** Selected clinical and cost inputs.

| **Quantity** | **Reference value** | **Basis** |
|---|---|---|
| Entry age, weight, and height | 50 years; 95 kg; 1.68 m (BMI 33.7 kg/m²) | Representative cohort meeting the criterion |
| Untreated annual diabetes hazard | 0.062 | Assumed for adults with BMI ≥ 30 and HbA1c 6.0% to 6.4% |
| Diabetes hazard ratio on 1 mg | 0.30 | SELECT higher-HbA1c subgroup (0.23), scaled on the log scale by the BELIEVE HbA1c response ratio (0.81) |
| Persistence | 65% at one year; 80% annually thereafter | Program assumptions; midyear discontinuation |
| Background mortality multiplier | 1.5 | Status First Nations all-cause mortality in BC, adjusted for a cohort aged 50 and older |
| Annual drug and dispensing cost | \$1,141.07 | 13 pens at $84.39; four $11 fees |
| Annual incremental monitoring | \$243 | Assumed at full exposure |
| Annual cost of diabetes | $2,726 Alberta; $1,646 Ontario-derived | Published diabetes state costs, inflated to projected 2026 dollars |

BMI = body mass index; HbA1c = hemoglobin A1c. The same clinical inputs were used with both diabetes cost sources. Supplement Table S5 and the code archive give the distributions and sources.

**Table 2.** Lifetime outcomes per person starting semaglutide.

| **Delay retained** | **Diabetes costs** | **QALYs gained (95% UI)** | **Cost difference (95% UI)** | **Cost per QALY** | **Probability cost-effective at \$50,000 per QALY** |
|---|---|---|---|---|---|
| 100% | Alberta | 0.131 (0.047 to 0.261) | −$38 (−$2,221 to \$2,436) | Cost-saving | 99.8% |
| 100% | Ontario-derived | 0.131 (0.047 to 0.261) | $1,633 (−$361 to \$4,265) | \$12,424 | 98.6% |
| 83% | Alberta | 0.124 (0.043 to 0.249) | $525 (−$1,455 to \$2,891) | \$4,250 | 99.3% |
| 83% | Ontario-derived | 0.124 (0.043 to 0.249) | $2,015 ($145 to \$4,613) | \$16,297 | 97.2% |
| 0% | Alberta | 0.084 (0.015 to 0.194) | $3,336 ($1,935 to \$5,499) | \$39,500 | 64.1% |
| 0% | Ontario-derived | 0.084 (0.015 to 0.194) | $3,916 ($2,327 to \$6,472) | \$46,376 | 53.0% |

QALY = quality-adjusted life-year; UI = uncertainty interval, the central 95% of paired probabilistic draws. Differences are semaglutide plus usual care minus usual care alone. The 83% label uses the unrounded STEP 1 ratio of diabetes-range prevalence contrasts.

**Figure 1.**
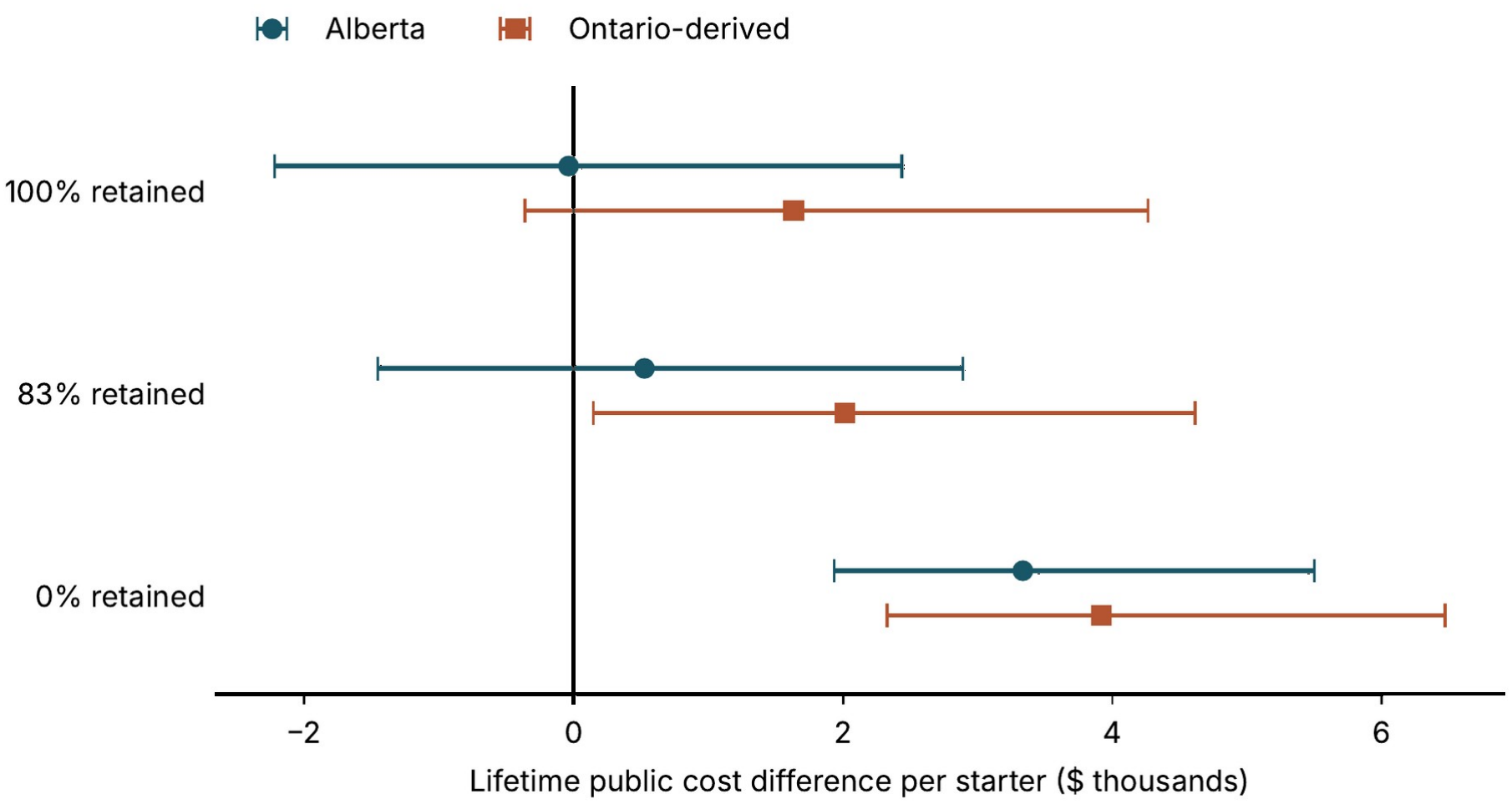
Lifetime incremental public costs per person starting semaglutide, by share of diabetes delay retained after stopping and by diabetes cost source. Points are mean incremental costs. Lines span the central 95% of draws within each scenario. Negative values indicate lower costs with semaglutide.

**Figure 2.**
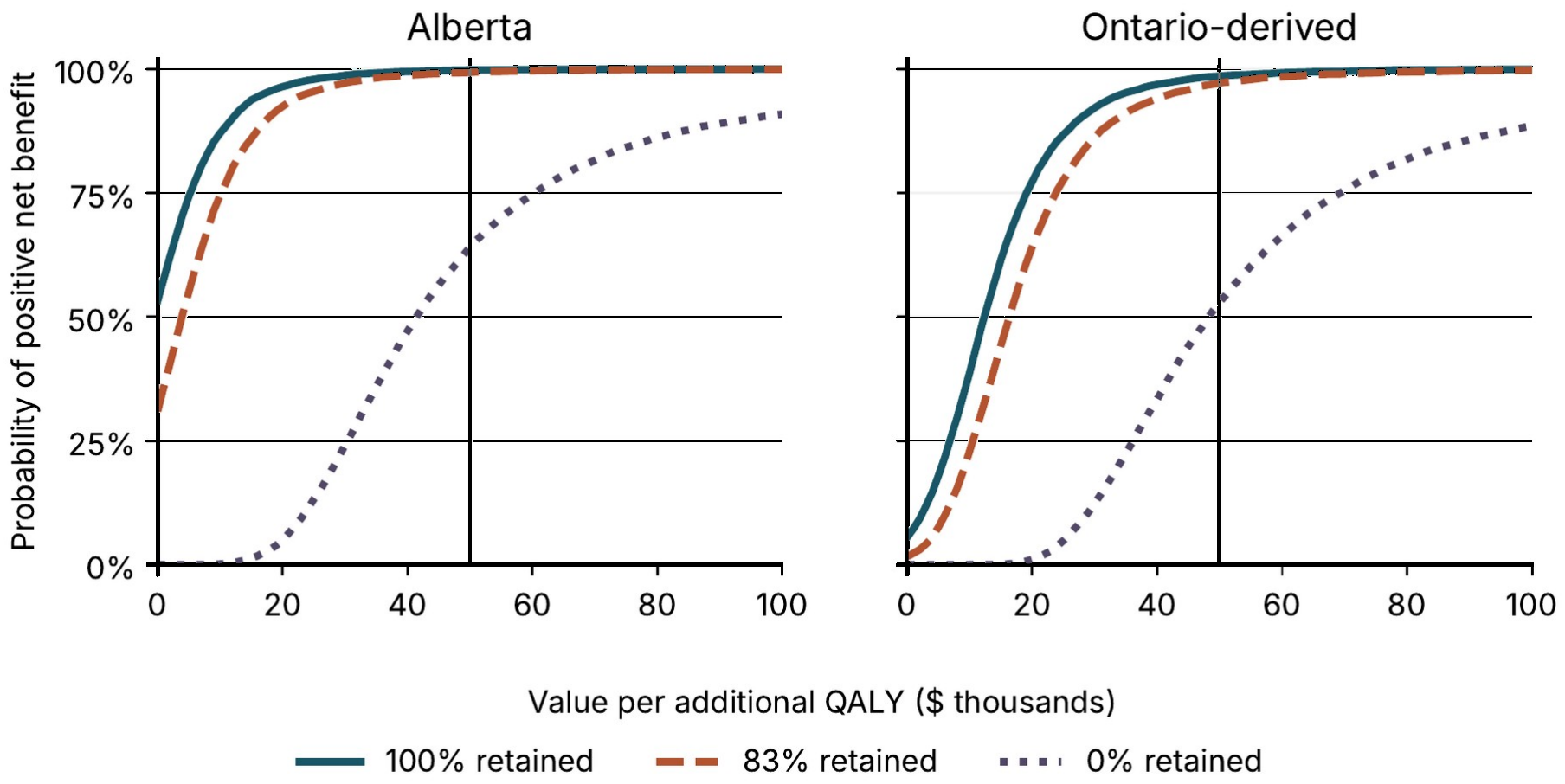
Probability that semaglutide is cost-effective, by value per quality-adjusted life-year (QALY). For each value per QALY, the curve shows the proportion of draws in which the value of the QALYs gained exceeds the additional cost. The vertical line marks $50,000 per QALY.

Using the STEP 1 diabetes-range contrast (83% retention), treatment gained 0.124 QALYs (95% UI 0.043 to 0.249). Mean public spending rose by $525 using Alberta costs (95% UI −$1,455 to $2,891) and by $2,015 using Ontario-derived costs (95% UI $145 to $4,613). The corresponding costs per QALY were $4,250 and $16,297, and the probabilities of positive net benefit at $50,000 per QALY were 99.3% and 97.2%. These gains reflect time spent without diabetes as well as the modelled effects on weight, cardiovascular events, and kidney failure.

With complete catch-up (0% retention), patients still benefited during treatment and during the year after stopping. Treatment gained 0.084 QALYs per person (95% UI 0.015 to 0.194). Spending rose by $3,336 using Alberta costs (95% UI $1,935 to $5,499) and by $3,916 using Ontario-derived costs (95% UI $2,327 to $6,472), yielding $39,500 and $46,376 per QALY. At $50,000 per QALY, the probability of positive net benefit was 64.1% with Alberta costs and 53.0% with Ontario-derived costs.

### Sensitivity analyses

One conservative analysis combined two unfavourable inputs: an untreated annual diabetes hazard of 0.03 and complete catch-up. Cost per QALY then rose to $55,231 using Alberta costs and $60,179 using Ontario-derived costs. Limiting new benefits to four years while continuing to charge for treatment increased the ratio under complete catch-up to $75,449 and $82,779. At 83% retention, the four-year limit yielded $24,767 per QALY using Alberta costs and $37,388 using Ontario-derived costs (Supplement Table S3). With the lower persistence seen in United States practice, costs and health gains both fell. Cost per QALY was $92 using Alberta costs and $11,965 using Ontario-derived costs at 83% retention, and $59,649 and $62,964 with complete catch-up.

At the Ontario comparator pen price, full retention yielded a mean saving of $336 per person using Alberta costs. At 83% retention, spending still increased by $227 using Alberta costs and $1,716 using Ontario-derived costs. The break-even pen prices at 83% retention were $73.38 using Alberta costs and $42.18 using Ontario-derived costs.

## Discussion

For Plan W adults with obesity and higher-risk prediabetes, the model projects that adding generic semaglutide to usual care would improve health at a modest additional lifetime cost. At the BC pen price and the STEP 1 estimate of the prevalence contrast, each additional QALY cost approximately $4,200 using Alberta costs or $16,300 using Ontario-derived costs. Only the combination of full retention and the higher annual diabetes cost yielded a mean saving at the reference inputs. That saving was small relative to the uncertainty around it.

The estimated health gains were identical for the two cost sources, so the difference in net spending comes from the annual cost of diabetes. At 83% retention, diabetes care avoided over a lifetime was worth $3,764 per person using the Alberta annual cost and $2,275 using the Ontario-derived annual cost. This $1,489 difference accounts for the difference in net spending. With full retention, the offsets grew to $4,224 and $2,552. With complete catch-up, less diabetes care was avoided, and the choice of cost source had a smaller effect (Figure 1).

The finding matters for a payer deciding whether to fund treatment before a patient has diabetes. Much of the projected return comes from postponing years of diabetes care. Even with complete catch-up after stopping, treatment had already provided some time without diabetes. Under that assumption, however, the Ontario-derived result was close to the illustrative benchmark, with almost equal probabilities of positive and negative net benefit. Lower untreated risk, lower persistence, or a shorter duration of benefit would make coverage less attractive. A local estimate of progression among untreated adults with HbA1c 6.0% to 6.4% would therefore materially improve the decision.

Withdrawal studies provide stronger evidence on weight regain than on the timing of diabetes. A patient can regain weight or move from normal HbA1c to prediabetes without developing diabetes. STEP 1 found that the semaglutide group still had a lower prevalence of diabetes-range HbA1c one year after stopping, but the contrast rested on five participants with diabetes-range values at follow-up. The 83% scenario is therefore best read as one point between the full-retention and complete catch-up bounds. In STEP 10, 28 weeks after treatment ended, diabetes was observed in 3 of 120 assessed participants assigned to semaglutide and 5 of 60 assigned to placebo. Neither study followed participants for more than one year after stopping, and neither supplies a post-withdrawal hazard for confirmed diabetes in this population. The model therefore extends a one-year observation over a lifetime. Measuring diagnoses and their timing would help distinguish a temporary rise after cessation from a return to ordinary risk.[18,23]

A Canadian evaluation by Rennert-May and colleagues reported $72,962 per QALY for semaglutide 2.4 mg in patients with established cardiovascular disease, rising to $190,465 when the duration of mortality benefit was limited to four years. Their decision concerned secondary prevention at a different dose and price. Here, a lower generic price and selection by HbA1c shift much of the value to diabetes prevention. Both evaluations are sensitive to how long benefits last.[24]

The annual diabetes costs deserve particular attention in BC. Plan W covers eligible prescriptions across adulthood, whereas some Ontario sources have incomplete capture of prescriptions for younger adults. Other BC plans, including Plan NP, may pay for diabetes medicines. Illustrative annual BC costs include approximately $313 for dapagliflozin, $1,182 for empagliflozin, and $547 to $702 for two insulin glargine products at 30 units daily, including dispensing. These are amounts per treated patient. Estimating an additional offset would require the prescribing mix, the payer’s net payments, and the share already represented in the published diabetes costs. [1,5,6,22,25]

Several limitations affect transfer to Plan W. The untreated diabetes hazard, persistence on treatment, and relative glycemic effect of the 1 mg dose have not been validated in adults meeting the criterion. The model captures mortality through selected cardiovascular and kidney pathways and may miss other consequences of diabetes. A broader mortality estimate would need to account for deaths already represented. The intervals describe uncertainty within each modelled course after withdrawal and assign no probabilities to those courses. Finally, the evaluation uses aggregate evidence. It does not establish patient preferences, local clinical priorities, or population uptake. Any linkage of dispensing, laboratory, and claims data would require appropriate First Nations governance and agreement about the use and interpretation of results.[16,26,27]

## Conclusions

Covering generic semaglutide for Plan W adults with BMI of 30 kg/m^2^ or more and HbA1c 6.0% to 6.4% could postpone diabetes and improve health at an acceptable additional cost. At the BC pen price and the reference inputs, the estimated cost per QALY ranged from cost-saving to $46,376 across the withdrawal courses and cost sources examined, and was $4,250 using Alberta diabetes costs and $16,297 using Ontario-derived costs when the model used the STEP 1 diabetes-range contrast. A payer could reasonably anticipate health gains from coverage. The evidence is less certain that treatment would pay for itself.

## Supporting information

Supplement: detailed methods, Tables S1 to S5 and Figure S1

Model code, parameter files and results

## Data Availability

This study used published aggregate data only. All model code, parameter files, and results produced in the present work are provided as supplementary files with this preprint.

## Ethics

This evaluation used published aggregate sources only. It involved no human participants and no individual-level records. The views expressed are those of the authors and do not represent the position of the Fraser Health Authority, Simon Fraser University, the University of British Columbia, the First Nations Health Authority, the Ministry of Health, or any First Nation.

## Data availability

The model code, parameter files, and results are provided as supplementary files with this preprint.

## Acknowledgments

Artificial intelligence-assisted tools (Claude, Anthropic; ChatGPT and Codex, OpenAI) were used under the authors’ direction to help write and check the model code, run and reproduce the analyses, and draft and edit the manuscript and supplement. The authors reviewed and verified the content and take responsibility for it.

