## Supplement: detailed methods, Tables S1 to S5 and Figure S1 for "Cost-effectiveness of generic semaglutide for Status First Nations adults with obesity and prediabetes under British Columbia’s Plan W"

### S1. Population, time horizon, and model structure

The cohort starts free of diagnosed diabetes and established cardiovascular disease at age 50, with a weight of 95 kg and height of 1.68 m (body mass index 33.7 kg/m²). In the code and data files this eligibility criterion is labelled C2b. The model updates five mutually exclusive event states in annual cycles: no event, post-myocardial infarction, post-stroke, kidney failure, and dead. Diabetes is tracked as a proportion of survivors. Event rates are converted to probabilities, costs and utilities follow midpoint occupancy, and follow-up ends at age 100. Annual costs and QALYs are discounted by 1/(1.015) raised to the midcycle time in years. The source files give the state transitions and parameter distributions.

Diabetes increases cardiovascular risk by an assumed factor of 1.5. The kidney failure rate of 9.3 per 10,000 diabetes person-years comes from Ontario First Nations data. Background mortality follows a Gompertz approximation to the Canadian life table, multiplied by 1.5 (range 1.2 to 1.85) to reflect higher all-cause mortality among Status First Nations people in BC. The published all-ages ratio was 1.85 in 2017. We used 1.5 for a cohort aged 50 and older because much of the excess occurs at younger ages. Cardiovascular deaths and mortality after kidney failure are represented separately. The model does not estimate all excess mortality attributable to diabetes. Neither a population-level First Nations diabetes incidence ratio nor a local calibration factor is applied to the diabetes hazard.

### S2. Diabetes and weight during and after treatment

SELECT measured a first HbA1c crossing at 6.5%. The 17.0% with diabetes-range HbA1c at week 156 describes status at a visit. The 778 first crossings among 2,794 placebo participants with higher HbA1c do not supply subgroup person-time. The untreated annual hazard of 0.062 is an assumption. It equals $-\ln(1 - 0.17)/3$, the constant rate that would give 17% with diabetes after three years. Lower and higher hazards were examined separately. The treatment hazard ratio at 1 mg was obtained from $\exp[0.81 \times \log(0.23)]$. The 0.81 scaling factor is the ratio of the placebo-adjusted HbA1c reductions with 1 mg and 2.4 mg in BELIEVE (0.25/0.31). It is applied on the log hazard scale as an inference. It is not the ratio of weight responses, which is lower (about 0.63).

On-treatment effects are scaled by the modelled proportion on treatment divided by 0.85, the assumed average on-treatment proportion within the SELECT intention-to-treat estimate, and capped at the trial effect.

#### Table S1. Diabetes-range HbA1c in the STEP 1 extension.

| Visit | Semaglutide 2.4 mg | Placebo | Placebo minus semaglutide |
|---|---|---|---|
| Week 68 | 0/141 (0%) | 3/53 (5.66%) | 5.66 percentage points |

| | | | |
|---|---|---|---|
| Week 120 | 2/120 (1.67%) | 3/47 (6.38%) | 4.72 percentage points |

Figure 3 of the STEP 1 extension classifies available participants with prediabetes at baseline. The counts and percentages with diabetes-range values are printed on the bars of that figure, and the denominators are consistent with the percentages reported in the text of the paper. The ratio of the two differences is (3/47 − 2/120) ÷ (3/53 − 0/141) = 0.833215. No individual transition matrix or off-treatment incidence hazard can be recovered from the bars. The 83% parameter is a structural extrapolation that rests on five participants with diabetes-range values at week 120. Ordinary progression and changing denominators both contribute to the observed contrast.

The model tracks the diabetes-free prevalence gap by time since stopping. Each discontinuing cohort carries its share of the gap into the next cycle. Usual diabetes risk resumes. In the first year after withdrawal, a multiplier declines linearly from one to its terminal value of one, 0.833215, or zero. Ordinary progression can narrow the gap in every scenario. The zero endpoint removes only the remaining gap for the cohort that stopped. It leaves past diabetes-free time, exposure, and benefit among continuing patients intact.

For the weight-related utility effect, STEP 1 gives a placebo-adjusted one-year residual of (5.6 − 0.1)/(17.3 − 2.0), or about 36% of the end-of-treatment difference. We used the same declining weight curve in every analysis and assigned no residual weight utility beyond the measured year. The weight curve does not determine the post-treatment diabetes hazard.

#### S3. Costs and prescription coverage

Thirteen four-dose pens at $84.39 plus four dispensing fees of $11 give annual acquisition and dispensing costs of $1,141.07. Drug costs, monitoring costs, and treatment effects accrue in proportion to exposure. The model does not separately reconstruct titration, early first fills, or wastage. For the lower-price test only the pen price changes. The four dispensing fees continue. The break-even price equals the BC price minus mean incremental cost divided by expected discounted pens.

Olivieri and colleagues report $2,352 for the Alberta diabetes input in 2021; Wiggins and colleagues use $1,459 for uncomplicated diabetes in 2022. Both were inflated using the annual Canadian health care CPI, projecting 2026 at 2% above 2025. The Alberta cost includes pharmacy under microvascular complications. The Ontario evidence includes model-derived estimates and sources with incomplete all-age prescription capture. Diabetes costs accrue outside the kidney failure state. Acute cardiovascular events are costed separately. No separate terminal-care or transport allowance is added without an independently costed service.

**Table S2. Examples of annual BC treatment costs after diabetes diagnosis.**

| Product | Daily regimen | Annual drug and dispensing cost |
|---|---|---|
| Dapagliflozin | 10 mg | $313 |
| Empagliflozin | 10 mg | $1,182 |
| Insulin glargine (Semglee) | 30 units | $547 |
| Insulin glargine (Basaglar) | 30 units | $702 |

These are illustrative full-year amounts including four dispensing fees. The two insulin costs use product-specific prices per mL. Actual spending depends on the drug, dose, timing, and public plan paying the claim. Plan NP covers listed diabetes medicines for eligible residents. Neither the propensity to prescribe these treatments nor their incremental public cost beyond the Alberta and Ontario inputs was measured here.

#### S4. Uncertainty and sensitivity analyses

We used 10,000 parameter draws with seed 2026. A draw contains uncertain clinical rates, treatment effects, costs, utilities, and persistence inputs. It is applied to both arms, and the same clinical draw is used for each cost source. Table S5 lists every sampled parameter that enters this analysis, with its reference value, range, distribution, realized draws, and basis. Figure S1 shows the sampled distributions. All other inputs are fixed and are listed in model/parameters.csv.

Each range is treated as an approximate 95% interval. For gamma, beta, and normal distributions the mean equals the reference value and the standard deviation equals the width of the range divided by 3.92. Where a range is not symmetric about the reference value, the sampled 2.5th and 97.5th percentiles therefore differ from the stated limits, as Table S5 shows. Hazard ratios and relative risks use a two-piece normal distribution on the log scale: the reference value is the median, and the lower and upper limits are the 2.5th and 97.5th percentiles, each with its own log-scale standard deviation. This keeps the published point estimate as the median when the interval is not symmetric on the log scale. The dose-scaling factor uses a triangular distribution with its mode at the reference value. Underlying inputs are sampled independently because joint estimates are unavailable. Derived quantities retain dependence on their shared inputs. No probabilities are assigned to the three withdrawal courses.

The lower and higher incidence tests scale the common hazard draws to point inputs of 0.03 and 0.10. The duration test stops new diabetes and weight-related benefits after model year four while treatment costs continue according to persistence. Prior diabetes delay follows the same withdrawal rules. The lower persistence test scales the persistence draws to 35.2% on treatment at one year and 44.3% annual retention thereafter, which reproduces the 64.8% and 84.4% discontinuation at one and two years reported for United States adults without type 2 diabetes by Rodriguez and colleagues. Results in Table S3 are ratios of incremental means.

**Table S3. Incremental cost per QALY under alternative clinical inputs.**

| Analysis | Delay retained | Alberta | Ontario-derived |
|---|---|---|---|
| Reference | 100% | Cost-saving | $12,424 |
| Reference | 83% | $4,250 | $16,297 |
| Reference | 0% | $39,500 | $46,376 |
| Lower incidence | 100% | $12,443 | $23,761 |
| Lower incidence | 83% | $17,527 | $28,088 |
| Lower incidence | 0% | $55,231 | $60,179 |
| Higher incidence | 100% | Cost-saving | $9,024 |

| | | | |
|---|---|---|---|
| Higher incidence | 83% | $142 | $12,448 |
| Higher incidence | 0% | $30,747 | $38,652 |
| Four-year benefit limit | 100% | $18,597 | $31,863 |
| Four-year benefit limit | 83% | $24,767 | $37,388 |
| Four-year benefit limit | 0% | $75,449 | $82,779 |
| Lower persistence | 100% | Cost-saving | $7,047 |
| Lower persistence | 83% | $92 | $11,965 |
| Lower persistence | 0% | $59,649 | $62,964 |

### Table S4. Lifetime costs at alternative pen prices.

| Delay retained | Diabetes costs | Cost at $84.39 | Cost at $78.141 | Break-even pen price |
|---|---|---|---|---|
| 100% | Alberta | −$38 | −$336 | $85.19 |
| 100% | Ontario-derived | $1,633 | $1,335 | $50.17 |
| 83% | Alberta | $525 | $227 | $73.38 |
| 83% | Ontario-derived | $2,015 | $1,716 | $42.18 |
| 0% | Alberta | $3,336 | $3,037 | $14.50 |
| 0% | Ontario-derived | $3,916 | $3,618 | $2.34 |

### Table S5. Parameters sampled in the probabilistic analysis.

| Group | Parameter | Reference value | Range | Distribution | Sampled median (2.5th to 97.5th percentile) | Basis |
|---|---|---|---|---|---|---|
| Clinical | Untreated annual diabetes hazard | 0.062 | 0.030 to 0.120 | Gamma | 0.059 (0.026 to 0.115) | Assumed. No observed estimate exists for this BMI and HbA1c group. Fixed alternatives of 0.03 and 0.10 are reported in Table S3. |
| Clinical | Hazard ratio for diabetes-range HbA1c, semaglutide 2.4 mg | 0.23 | 0.19 to 0.26 | Lognormal, two-piece | 0.23 (0.19 to 0.26) | SELECT, higher-HbA1c subgroup (Kahn 2024). Published point estimate and 95% confidence interval. |
| Clinical | Share of the 2.4 mg log hazard reduction applied at 1 mg | 0.81 | 0.63 to 0.95 | Triangular | 0.80 (0.67 to 0.92) | BELIEVE placebo-adjusted HbA1c reductions with 1 mg and 2.4 mg (0.25/0.31). Upper limit set below 1 so that the doses are never treated as equivalent. |
| Clinical | Proportion on treatment at one | 0.65 | 0.47 to 0.80 | Beta | 0.65 (0.48 to 0.80) | Assumed from published |

| | | | | | | |
|---|---|---|---|---|---|---|
| | year | | | | | persistence studies (Rodriguez 2025 and others). Not measured in Plan W. |
| Clinical | Annual retention after year one | 0.80 | 0.65 to 0.90 | Beta | 0.81 (0.66 to 0.91) | Assumed. No Canadian data. |
| Clinical | Weight loss with semaglutide 1 mg, proportion of body weight | 0.098 | 0.085 to 0.111 | Normal, truncated at 0 | 0.098 (0.085 to 0.111) | BELIEVE, 1 mg arm. |
| Clinical | Weight loss with usual care, proportion of body weight | 0.035 | 0.024 to 0.046 | Normal, truncated at 0 | 0.035 (0.024 to 0.046) | BELIEVE, placebo arm with lifestyle counselling. |
| Clinical | Major cardiovascular events per 100 person-years without diabetes | 0.70 | 0.40 to 1.20 | Gamma | 0.68 (0.35 to 1.15) | Assumed for adults aged about 50 with BMI ≥ 30 and prediabetes. |
| Clinical | Relative cardiovascular event rate after diabetes | 1.50 | 1.20 to 2.00 | Lognormal, two-piece | 1.50 (1.21 to 1.99) | Assumed. |
| Clinical | Kidney failure per 100 person-years with diabetes | 0.093 | 0.047 to 0.130 | Gamma | 0.092 (0.056 to 0.138) | Ontario First Nations cohort (Nash 2019), 9.3 per 10,000 person-years. |
| Clinical | Annual mortality with kidney failure | 0.15 | 0.10 to 0.20 | Beta | 0.15 (0.10 to 0.20) | Assumed. |
| Clinical | Background mortality multiplier | 1.50 | 1.20 to 1.85 | Lognormal, two-piece | 1.50 (1.20 to 1.86) | BC Status First Nations all-cause age-standardized mortality ratio of 1.85 in 2017 (First Nations Health Authority and Provincial Health Officer 2024). A value of 1.5 was used for a cohort aged 50 and older because much of the excess occurs at younger ages. |
| Utility | Baseline utility | 0.80 | 0.75 to 0.85 | Beta | 0.80 (0.75 to 0.85) | Assumed for adults with obesity without diabetes. |
| Utility | Utility gain per unit of BMI lost | 0.0061 | 0.0000 to 0.0122 | Normal, truncated at 0 | 0.0061 (0.0001 to 0.0123) | Value used in Beaudet 2014. Lower limit of zero. |
| Utility | Gastrointestinal adverse-effect disutility, year one | 0.010 | 0.000 to 0.030 | Gamma | 0.008 (0.001 to 0.029) | Assumed. |
| Utility | Disutility of diabetes | 0.020 | 0.000 to 0.050 | Gamma | 0.017 (0.003 to 0.052) | Assumed for uncomplicated type 2 diabetes. |
| Utility | Disutility after myocardial | 0.055 | 0.030 to 0.080 | Gamma | 0.054 (0.033 to 0.083) | Beaudet 2014. |

| | | | | | | |
|---|---|---|---|---|---|---|
| | infarction | | | | | |
| Utility | Disutility after stroke | 0.164 | 0.100 to 0.220 | Gamma | 0.162 (0.109 to 0.231) | Beaudet 2014. |
| Utility | Disutility of kidney failure | 0.164 | 0.050 to 0.250 | Gamma | 0.158 (0.079 to 0.281) | Beaudet 2014. |
| Cost | Annual cost of diabetes, Alberta | $2,726 | $1,800 to $3,500 | Gamma | $2,701 ($1,936 to $3,650) | Olivieri 2024, inflated to 2026. |
| Cost | Annual cost of diabetes, Ontario-derived | $1,646 | $800 to $2,400 | Gamma | $1,615 ($936 to $2,550) | Wiggins 2025, inflated to 2026. |
| Cost | Annual incremental monitoring on treatment | $243 | $0 to $350 | Gamma | $233 ($101 to $446) | Four physician visits and two laboratory panels, the resource assumption in Olivieri 2024. |
| Cost | Annual treatment-related hospital cost on treatment | $15 | $0 to $100 | Gamma | $5 ($0 to $89) | Excess gallbladder events in SELECT multiplied by a BC cholecystectomy cost. |
| Cost | Myocardial infarction, acute event | $11,963 | $8,000 to $16,000 | Gamma | $11,854 ($8,372 to $16,243) | Canadian Institute for Health Information Patient Cost Estimator, BC, inflated. |
| Cost | Myocardial infarction, annual follow-up | $1,739 | $1,000 to $3,000 | Gamma | $1,697 ($895 to $2,863) | Olivieri 2024 supplement, inflated. |
| Cost | Stroke, first-year attributable cost | $38,859 | $23,987 to $44,976 | Gamma | $38,647 ($29,110 to $49,846) | Vyas 2023, inflated. |
| Cost | Stroke, annual follow-up | $4,508 | $2,500 to $6,000 | Gamma | $4,443 ($2,923 to $6,388) | Olivieri 2024 supplement, inflated. |
| Cost | Kidney failure with dialysis, annual | $111,521 | $80,000 to $130,000 | Gamma | $111,150 ($87,953 to $137,896) | Alberta one-year cost reported by Manns and colleagues (2019), inflated. |
| Cost | Unrelated public health care cost per year alive | $4,000 | $3,000 to $6,000 | Gamma | $3,942 ($2,638 to $5,655) | Assumed. |

The reference value is the mean of the gamma, beta, and normal distributions and the median of the lognormal distributions. Sampled values are from the 10,000 draws used in the analysis. Full source notes for each parameter are in model/parameters.csv.

**Figure S1. Sampled distributions of the parameters in Table S5.**

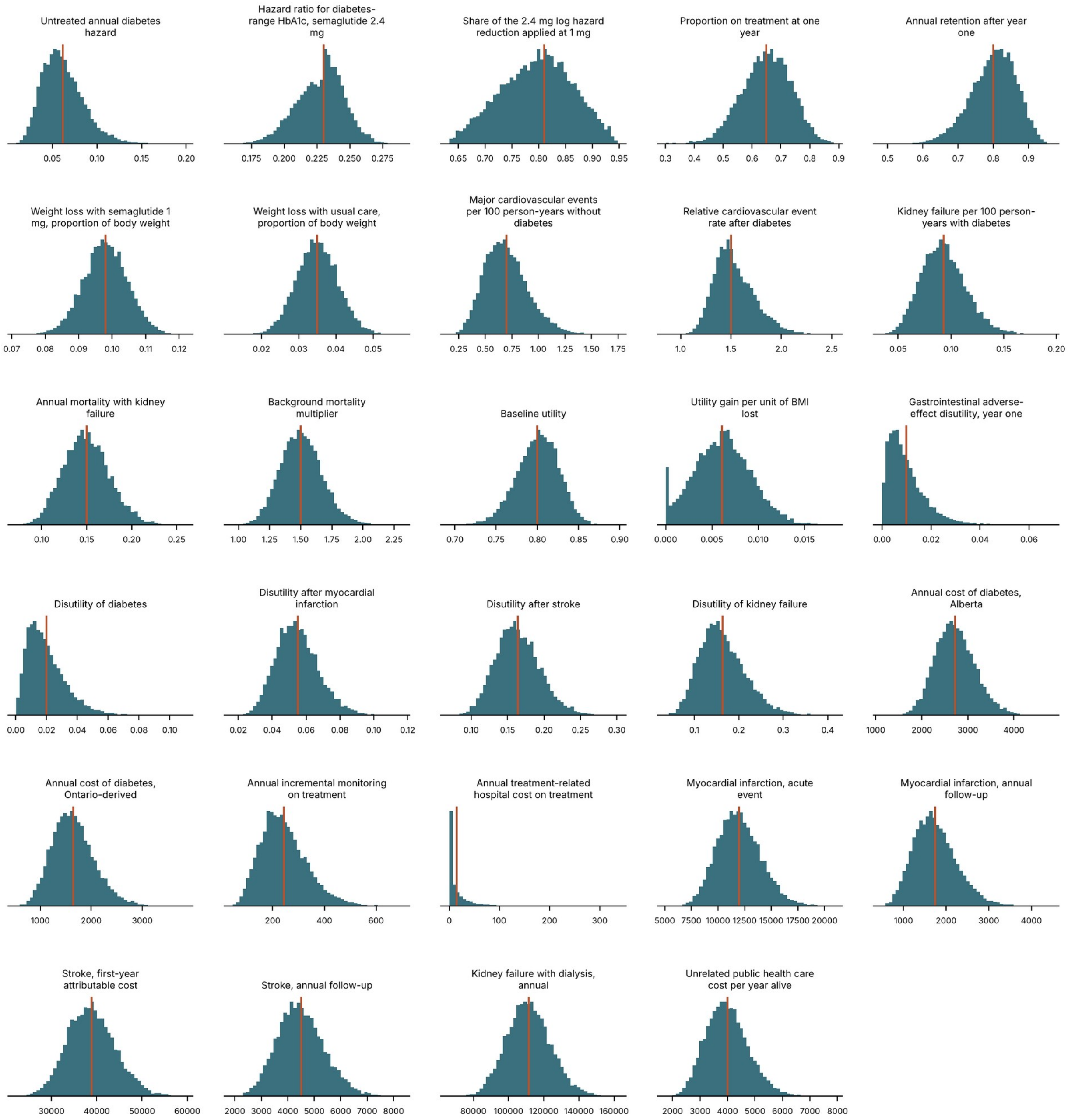

Each panel shows the 10,000 draws for one parameter. The vertical line marks the reference value.

### S5. Analysis files and sources

The archive contains parameter values and sources, model equations, and executable analysis and plotting scripts written in Python with NumPy, pandas, and Matplotlib. Data.csv gives the three reference scenarios and the four clinical sensitivity analyses using both cost specifications. The results directory also contains arm-level estimates and the points underlying the cost-effectiveness acceptability curves. The analysis used published aggregate data. There was no prospectively registered analysis protocol or patient-level resampling.

The clinical evidence, costing, and modelling sources are listed in the references of the main paper. Briggs and Sculpher introduce cohort models. Further technical guidance is given by Siebert U, Alagoz O, Bayoumi AM, et al. State-transition modeling: A report of the ISPOR-SMDM Modeling Good Research Practices Task Force-3. Value Health 2012;15:812-820. doi:10.1016/j.jval.2012.06.014
